# Integration of serum androgens and Sex Hormone-Binding Globulin for optimized early detection of aggressive prostate cancer

**DOI:** 10.1101/2024.12.05.24318544

**Authors:** Olga Lazareva, Anja Riediger, Oliver Stegle, Holger Sültmann, Markus Hohenfellner, Magdalena Görtz

**Affiliations:** Junior clinical cooperation unit ‘Multiparametric Methods for Early Detection of Prostate Cancer’, German Cancer Research Center (DKFZ), Heidelberg, Germany; Division of Computational Genomics and Systems Genetics, German Cancer Research Center (DKFZ), Heidelberg, Germany; European Molecular Biology Laboratory, Genome Biology Unit, Heidelberg, Germany; Wellcome Sanger Institute, Wellcome Genome Campus, Hinxton, UK; Department of Urology, University Hospital Heidelberg, Heidelberg, Germany; National Center for Tumor Diseases (NCT), Heidelberg, Germany; Faculty of Biosciences, Heidelberg University, Heidelberg, Germany; Division of Cancer Genome Research, German Cancer Consortium (DKTK), German Cancer Research Center (DKFZ), Heidelberg, Germany

**Keywords:** Androstenedione, Dehydroepiandrosterone-Sulfate, Early Detection of Cancer, Prostatic Neoplasms, Prostate-Specific Antigen, Risk Assessment, Sex Hormone-Binding Globulin

## Abstract

**Background:** Aggressive prostate cancer (PC) represents a significant health concern worldwide. Conventional initial screening methods, primarily based on prostate-specific antigen (PSA) levels, lack specificity, leading to a high rate of unnecessary biopsies and an urgent need for more accurate diagnostic tools. This study addresses the gap by exploring the potential of integrating clinical and routine blood laboratory parameters including a comprehensive hormone assessment to enhance the non-invasive prediction of aggressive PC.

**Methods:** In a pilot study of 578 patients who were scheduled for a prostate biopsy due to suspicion of PC, we analyzed an extensive panel of 28 laboratory values alongside data on family history, diet, and lifestyle. A logistic regression classifier was developed, and model performance was evaluated using repeated k-fold cross-validation on the complete dataset (n=282). Participants were histologically categorized into three risk groups: healthy, moderate PC (ISUP 1-2 PC), and aggressive PC (ISUP 3-5 PC).

**Results:** Significant associations were found between PC aggressiveness and lower levels of androstenedione, Dehydroepiandrosterone-Sulfate (DHEA-S) and free PSA percentage, as well as higher levels of Sex Hormone Binding Globulin (SHBG). The integration of these serum markers with clinical parameters into a new multi-stage risk classifier for PC prediction significantly improved the predictive accuracy. The risk model outperformed PSA-only methods, demonstrating higher sensitivity and specificity in predicting aggressive PC.

**Conclusions:** Incorporating serum markers DHEA-S, androstenedione, and SHBG into a novel risk classifier can improve early detection of aggressive PC. These widely available and cost-effective blood biomarkers could reduce reliance on invasive prostate biopsies and expensive magnetic resonance imaging by providing a more targeted approach to non-invasive prediction of aggressive PC following PSA testing. Our pilot study lays the groundwork for larger-scale research to further explore the integration of androgens and SHBG in future risk stratification models for improved clinical decision making.

## INTRODUCTION

Prostate cancer (PC) is the second most prevalent cancer in men, representing 27% of all new cancer diagnoses in males (1). It is a major cause of mortality worldwide, with projections of the Lancet Commission on PC indicating a doubling of new cases between 2020 and 2040, particularly in low-income countries (2). Early diagnosis through screening can offer numerous benefits such as a higher likelihood of cure, less aggressive treatment options, reduced progression to metastatic stages, and improved quality of life (3–5). Prostate-specific antigen (PSA) screening is a widely used method for early detection of PC. Recent results from the Rotterdam Section of the European Randomized Study of Screening for PC indicate that PSA screening can substantially reduce long-term mortality rates (6). However, PSA testing has limited specificity, frequently resulting in unnecessary biopsies and the detection of indolent PC (7).

To address the limitations of PSA testing, the American Urological Association (AUA) guidelines recommend integrating risk calculators into the traditional decision-making process for PC screening (8). The primary tool is the PSA blood test, and for cases with elevated PSA levels, the AUA guidelines advise using risk calculators that include PSA, free PSA %, age, ethnicity, and family history for personalized risk assessment. Clinicians are then encouraged to engage in shared decision-making with patients, discussing the value of additional diagnostic data from biomarkers or multiparametric magnetic resonance imaging (mpMRI) and considering the advisability of proceeding with a biopsy. Similarly, the European Association of Urology (EAU) recommends using risk stratification nomograms for patients with elevated PSA, including age, family history, digital rectal examination (DRE), prostate volume, and PSA density (PSAD) (9). This method helps to identify men at low risk who may only require clinical follow-up, thereby reducing the need for expensive or invasive tests such as mpMRI and biopsy.

mpMRI and fusion-targeted biopsy of suspicious mpMRI lesions are advanced diagnostic methods for PC, significantly enhancing detection accuracy (10,11). High cost and restricted availability of mpMRI, especially in low-resource settings, limit its routine use as a screening tool (12). The economic and logistical challenges of MRI highlight the need for cost-effective and accessible biomarkers to facilitate broader implementation across diverse healthcare settings, improving early detection capabilities.

Recent research increasingly focuses on modifiable risk factors, such as lifestyle and diet, associated with PC risk (13,14). Metabolic changes linked to many diseases, including cancer, are reflected in human blood metabolomic patterns, which provide valuable insights into underlying physiological states (15). Systemic information from blood parameters, such as C-reactive protein, amino acids, and glycated hemoglobin, has been predictive of multiple cancers (16). Additionally, adrenal androgens influence the aetiopathogenesis of PC and are associated with aggressive PC (17),(18).

The study aimed to improve non-invasive prediction of aggressive PC by integrating 44 laboratory and clinical parameters into a predictive model. Our findings demonstrate that incorporating these parameters after PSA testing significantly improved diagnostic accuracy. This approach could reduce unnecessary biopsies, decrease the number of costly MRIs, and more accurately predict PC aggressiveness, aligning with precision medicine principles. By validating a multifaceted diagnostic strategy over traditional PSA-only methods, our research supports a more nuanced approach to PC screening and decision-making.

## MATERIALS AND METHODS

### Study population

565 men who received a prostate biopsy at the University Hospital Heidelberg between June 2021 and December 2023 were recruited for supplementary clinical and, if a preoperative blood sample had to be taken and patient consent was given, laboratory values assessment. They were eligible for study inclusion in case of suspicion of PC with a PSA level ≥3 ng/ml and/or suspicious DRE and no previous treatment for PC. Additionally, 13 healthy men with a PSA <2.5 ng/ml who presented to the Urology Clinic at Heidelberg University Hospital for benign conditions, were recruited to enlarge the healthy control group. Data was collected prospectively, and institutional review board approval was obtained (S-130/2021). Clinical information regarding family history (defined as having a first-degree relative with PC), lifestyle, etc. was assessed via questionnaire. Laboratory parameters included, among others, lipid metabolism, inflammatory markers, hormones, and vitamins (**Supplementary Table 1**). After obtaining informed consent, blood samples were collected via vein puncture between 8 and 11 AM in fasting patients. All blood samples were analyzed in the central laboratory of Heidelberg University Hospital, in addition to the standard preoperative lab values. The central laboratory of Heidelberg University Hospital has been accredited according to DIN EN ISO 15189 since 2005.

### Prostate biopsy

Prebiopsy mpMRI was performed on 543 patients with suspicion of PC and eligibility for mpMRI. All mpMRI scans followed the PI-RADS recommendations, according to European Society of Urogenital Radiology guidelines (19). Men underwent transperineal fusion-targeted biopsy using a UroNav system (Philips Invivo, Gainsville, FL, USA) of MRI-suspicious lesions and saturation biopsy adjusted to prostate volume, as previously described (20,21). Histopathological analyses were performed according to International Society of Urological Pathology (ISUP) standards. The participants were categorized into three risk groups: healthy patients (PSA <2.5 ng/ml or no PC in biopsy), moderate PC (ISUP 1-2 PC), and aggressive PC (ISUP 3-5 PC)(10).

### Statistical analysis

In this study, 28 laboratory and 16 clinical parameters were analyzed to determine their impact on PC risk. Each parameter was initially tested for its ability to discriminate between the three predefined risk groups: healthy patients (n=253), moderate PC (n=229), and aggressive PC (n=96) through statistical analysis. For continuous variables, an ordinary least squares (OLS) methodology was employed, fitting models for each parameter to predict the risk groups and subsequently testing the significance of the slope coefficients 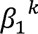:

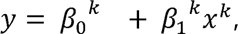

where *y* ∈ {0,1,2} is a risk category corresponding to healthy patients, ISUP 1-2 and ISUP 3-5 PC, 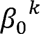 and 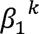 are intercept and slope coefficients for the *k*-th tested parameter *x^k^*.

For categorical variables the Chi-square test was used (22). All p-values were reported and subsequently adjusted for multiple comparisons using the Benjamini & Hochberg correction (23) with a family-wise error rate at 0.1 level. Complete results are provided in **Supplementary Table 1**, with significant findings highlighted in **Figure 1**.

**Figure 1:**
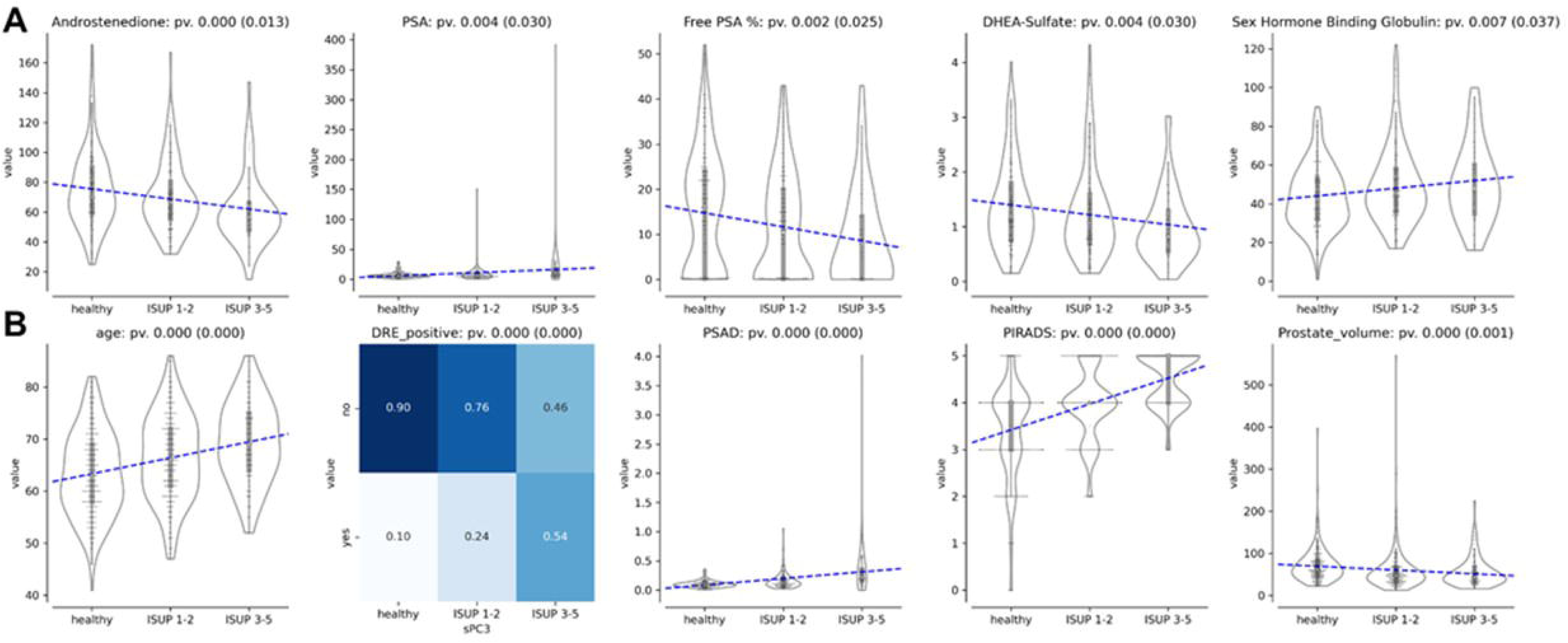
Statistically significant associations with the presence of moderate or aggressive PC. **A** Statistically significant association between laboratory parameters and the presence of ISUP 1-2 PC, ISUP 3-5 PC, or healthy patients. **B** Statistically significant associations between clinical and imaging parameters and the presence of ISUP 1-2 PC, ISUP 3-5 PC or healthy patients.

### Risk Modeling

Risk modeling was performed on 282 patients who had available data for clinical and laboratory parameters, showing a significant association with ISUP grade. The logistic regression model was developed in a staged approach:

- 1. Baseline model: Initiated with PSA, the standard marker for PC screening, to establish a benchmark for model performance.
- 2. Serum marker integration: Added laboratory measurements that showed a statistically significant association with PC risk, including Androstenedione, free PSA %, Dehydroepiandrosterone-sulfate (DHEA-Sulfate), and Sex Hormone Binding Globulin (SHBG) to evaluate improvements in sensitivity and specificity.
- 3. Clinical parameter integration: Included clinical factors from international guidelines (age, family history, DRE results, prostate volume, and PSAD) to assess their combined predictive value.
- 4. MRI results: Added PIRADS scores to capture additional risk information provided by imaging.
- 5. Advanced baseline model: Combined MRI and PSA.

Model performance was assessed at each stage by calculating the F1 score, Precision, Recall, and Accuracy. The model was evaluated using a repeated k-fold cross-validation (*k* = 10, repeated 10 times) approach, and all performance metrics were reported based on the test set (n=282). We used ‘L2’ regularization to decrease overfitting probability with the default regularization strength (*λ* = 1) using scikit-learn models (24).

### Interpretative Analysis Using SHAP Values

To understand the influence of individual variables on predictive outcomes, SHAP (SHapley Additive exPlanations) methodology was applied, following the approach proposed by Lundberg and Lee (25). This method quantitatively measured the impact of each parameter included in the model, providing clear insights into how specific factors contribute to overall risk prediction.

## RESULTS

### Patient cohort description

The study included a total of 578 patients. During the initial phase of the analysis, when statistically significant markers were selected, all available measurements were used without restricting to a fully overlapping cohort, maximizing the use of the data. Each laboratory parameter was represented by a minimum of 314 and a maximum of 353 measurements, while other parameters ranged from 314 to 578 measurements. A comprehensive list of these parameters and their respective counts is provided in **Supplementary Table 1**.

Following feature selection, the cohort was refined to include only patients with complete data across all parameters identified as significant (post-multiple hypothesis correction). This subset comprised 282 patients. Detailed descriptions of these patients and their corresponding data are presented in **Table 1**.

**Table 1.** Patient characteristics (modeling cohort n = 282) in the healthy, ISUP 1-2 PC vs. ISUP 3-5 PC groups for significant parameters. For continuous parameters, median and IQR are provided, otherwise, count per group and percentage are shown.

| Parameter | Unit | Healthy patients (n=122) | ISUP 1-2 PC (n=117) | ISUP 3-5 PC (n=43) |
| --- | --- | --- | --- | --- |
| Age | Years | 63 (57-69) | 66 (61-73) | 72 (64.50-75) |
| PSA | ng/dl | 5.75 (4.47-8.06) | 5.81 (4.2-8.41) | 10.11 (6.67-15.49) |
| Androstenedione | µg/l | 70 (59-88) | 66 (53-78) | 58 (48-67) |
| Free PSA % | % | 16 (0.30-25) | 11 (0.24-20) | 8 (0.18-17.50) |
| DHEA-Sulfate | µg/ml | 1.23 (0.76-1.75) | 1.06 (0.69-1.47) | 0.76 (0.54-1.14) |
| Sex Hormone Binding Globulin | nmol/l | 43 (32-53) | 46 (35-58) | 48 (35.5-62) |
| Prostate volume | ml | 61.32 (45-81.97) | 46.37 (34-66.45) | 45 (33.94-60) |
| PSAD | ng/ml/ml | 0.1 (0.07-0.13) | 0.13 (0.08-0.19) | 0.21(0.16-0.35) |
| Positive family history | Count, % | 12 (9.83) | 14 (11.97) | 7 (16.28) |
| Positive DRE | Count, % | 10 (8.20) | 72 (61.54) | 21 (48.84) |
| PIRADS ≤2 | Count, % | 19 (15.57) | 6 (5.13) | 0 (0) |
| PIRADS 3 | Count, % | 44 (36.07) | 23 (19.66) | 1 (2.33) |
| PIRADS 4 | Count, % | 53 (43.44) | 58 (49.57) | 14 (32.56) |
| PIRADS 5 | Count, % | 6 (4.92) | 30 (25.64) | 28 (65.12) |

### Role of blood biomarkers in prostate cancer

Significant associations were found between hormonal markers and PC severity as classified by ISUP grades (**Figure 1**). Lower levels of androstenedione and DHEA-Sulfate were significantly associated with more aggressive forms of PC (ISUP grades 3-5) (corrected p-values 0.013 and 0.03, respectively). This finding supports the hypothesis that reductions in certain androgens may signal advanced disease.

The bioavailability of circulating androgens is largely determined by the concentration of SHBG. Accordingly, in our study higher levels of SHBG were associated with increased PC severity (corrected p-value 0.037).

Higher PSA levels were positively correlated with PC severity (corrected p-value 0.03). Additionally, a lower percentage of free PSA % indicated more aggressive cancer (corrected p-value 0.025). These results reinforce the role of PSA and free PSA percentage as critical biomarkers in assessing PC aggressiveness.

### Incorporating clinical and lab values into new risk models for non-invasive prostate cancer prediction

In developing a PC risk classifier, we included the five significant laboratory values, along with clinical features and imaging parameters, that were strongly associated with ISUP grades. Age, DRE results, PSAD, PIRADS scores, and prostate volume showed statistically significant associations with ISUP grade severity, all with corrected p-values <0.05. Although a family history of PC did not show a significant statistical association with ISUP grades (corrected p-value 0.552), it was included in the model to adhere to AUA and EAU guidelines (8,9).

The performance of the risk classifier was evaluated through a multi-stage approach:

1. Baseline model initiated with PSA, 2. Enhancement with laboratory biomarkers significantly associated with PC risk (Androstenedione, free PSA %, DHEA-Sulfate, and SHBG), 3. Integration of clinical factors (Age, family history, DRE results, prostate volume, and PSAD), 4. Integration of MRI results (PIRADS scores) and 5. Advanced baseline model with solely PSA and MRI as comparison. This stepwise model enhancement resulted in gradual improvements across all metrics from stage 1-4, including F1 score, precision, recall, and accuracy. The progression of median scores through each stage is depicted in **Figure 2A**, with exact values provided in **Supplementary Table 2**.

**Figure 2:**
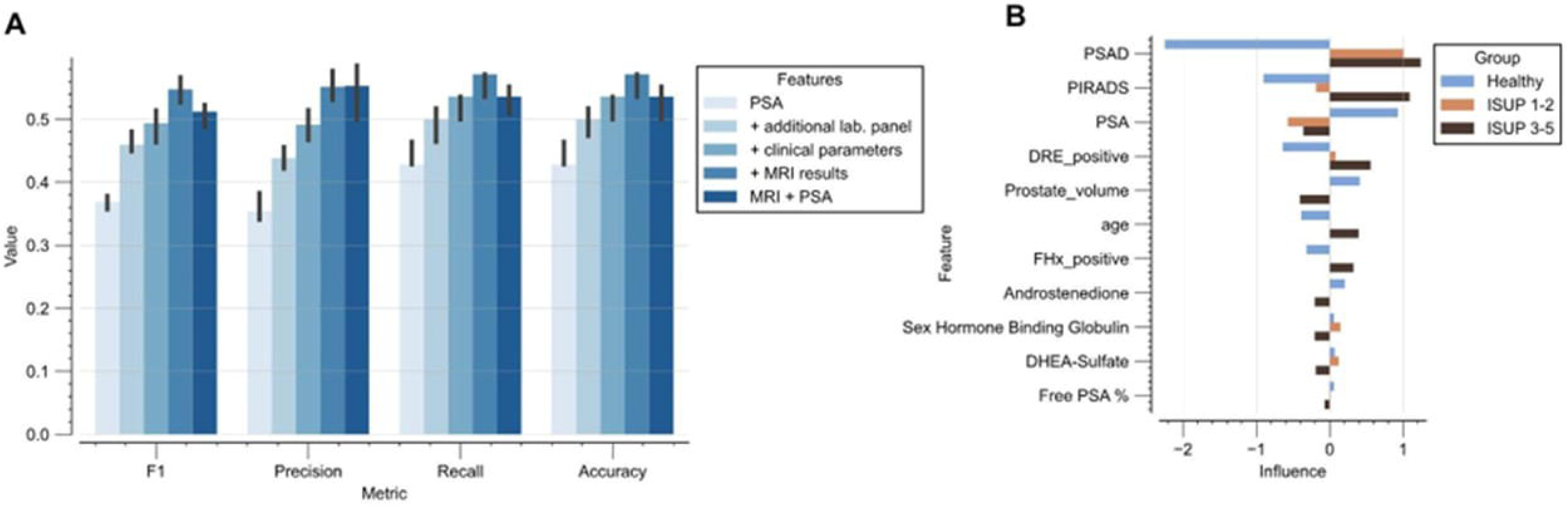
Risk modeling of PC. **A** Performance metric of a logistic classifier based on five sets of features: 1. PSA, 2. Integration of additional laboratory biomarkers, 3. Integration of clinical factors, 4. Integration of MRI results and 5. Solely PSA and MRI as comparison. **B** Feature importance for the three groups (healthy patients, ISUP 1-2 PC and ISUP 3-5 PC).

The regression coefficient distribution was analyzed to assess the prognostic significance of each parameter (**Figure 2B**). PSAD emerged as a particularly influential factor in identifying healthy patients. Established diagnostic parameters like PSA, DRE results and PIRADS also showed high importance for non-invasive prediction of PC presence. However, with these parameters, coefficients for ISUP grades 1-2 and 3-5 PC often shared the same sign, indicating challenges in differentiating these two categories. Serum androgens displayed variable signs for ISUP 1-2 versus ISUP 3-5 PC, suggesting their potential impact on the model’s predictive capability.

### Influence of individual variables on predictive outcomes using SHAP Values

To understand the influence of individual variables on predictive outcomes for each patient, we employed SHAP analysis to obtain per-patient SHAP values. These values help to identify the specific factors contributing to each patient’s prediction (PC versus no PC) and provide insight into the influence of each variable on the predictive outcomes. The model was developed to assist urologists in making informed decisions regarding prostate biopsies, especially when the complexity of individual patient profiles challenges clinical judgement. Values from two study patients are shown where biopsies didn’t reveal PC, and the model correctly predicted the absence of PC, demonstrating the practical utility of SHAP analysis as a transparent, data-driven foundation for clinicians **(Figure 3A and 3B)**.

**Figure 3:**
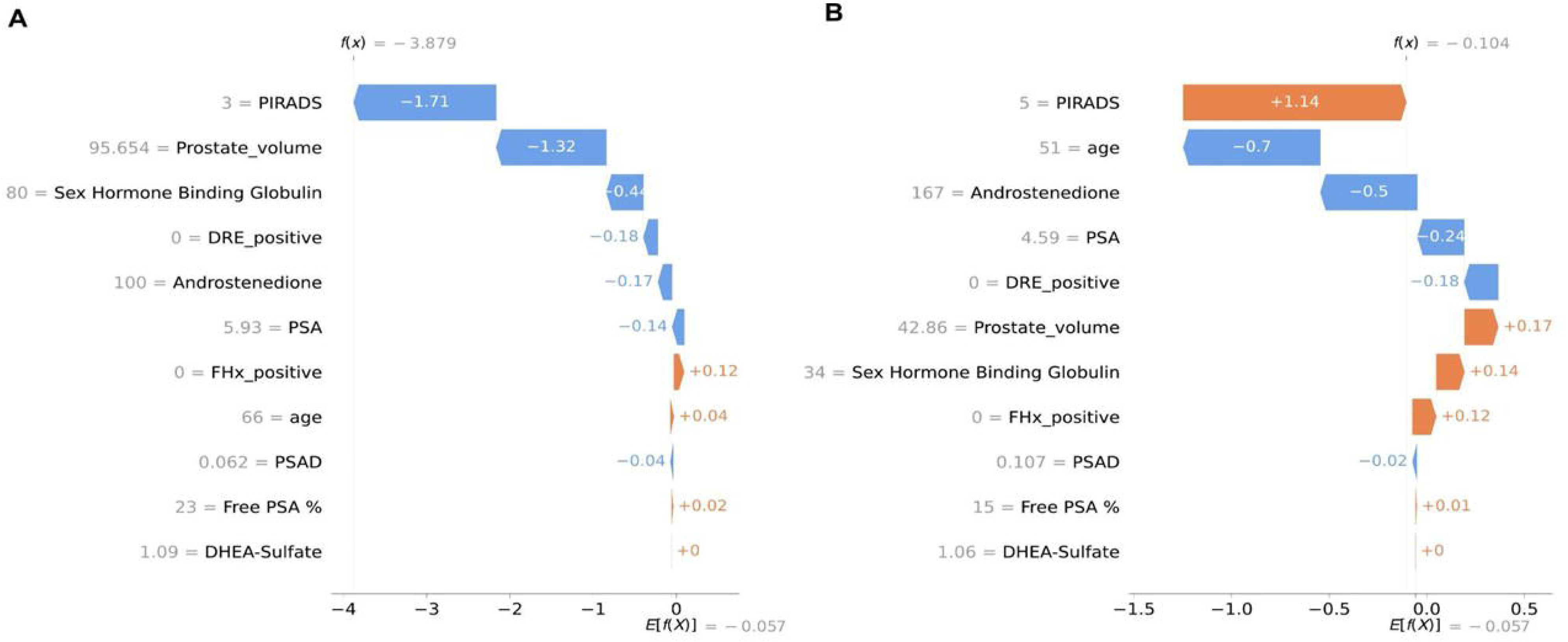
SHAP analysis to obtain per-patient SHAP values. SHAP values for two patients of the cohort who had no PC in the prostate biopsy: blue color indicates a reduction of the probability of having PC, and orange color indicates an increased probability of having PC.

## DISCUSSION

This study aimed to improve the non-invasive prediction of aggressive PC by integrating clinical and laboratory parameters into a comprehensive risk model. By identifying novel, cost-effective serum markers, we sought to improve decision-making following PSA testing and reduce reliance on invasive procedures and expensive MRI use.

Our study revealed several results. First, levels of DHEA sulfate and androstenedione were significantly reduced in newly diagnosed aggressive PC compared to healthy controls. In line with these results, higher levels of SHBG were significantly associated with increased PC aggressiveness. These markers had a stronger association with aggressive PC detection than established variables advised for use in risk stratification nomograms, such as family history (8,9). Thus, androgens have potential as diagnostic markers for early detection of aggressive PC, offering a complementary diagnostic tool alongside PSA testing. Androgens were particularly effective in distinguishing between ISUP 1-2 and ISUP 3-5 PC, highlighting their potential in differentiating moderate from aggressive PC, e.g. for treatment approaches such as active surveillance (26). ISUP 1-2 PC patients may be offered active surveillance instead of radical therapy due to the indolent nature of their disease (27). Existing biomarkers expressed by all PC, including indolent tumors, offer limited potential to selectively detect high-grade disease (28). Novel biomarkers distinguishing high-grade from low-grade PC are urgently needed, aligning with guidelines emphasizing a focus on high-grade PC due to the indolent nature of low-grade PC (8).

Second, integrating clinical findings and laboratory values into the decision-making process for recommending MRI scans has the potential to improve the selection of patients who benefit most from imaging. MRI remains time- and resource-intensive and may pose considerable costs for patients, apart from limitations in availability and inter-rater reliability. Barriers in healthcare delivery limit the wide adoption of MRI in real-world practice as a diagnostic test following PSA. The decision to use MRI and fusion biopsy can be supported by relatively inexpensive routine blood values and clinical parameters, minimizing unnecessary MRIs for PSA-positive patients with a low likelihood of PC. Commercially available blood values are straightforward to implement and less resource-intensive, allowing urologists to more selectively apply MRI and/or biopsy (29,30). Our results support implementing integrated decision-making frameworks in clinical routine to optimize MRI and biopsy use in PC diagnostics.

Third, the application of an explainable decision support concept in our study demonstrates an approach to guide urologists and patients in making informed decisions regarding prostate biopsies. A transparent and comprehensive assessment of each patient’s clinical scenario, including evaluation of clinical, lab values, and, if applicable, MRI findings, allows urologists and patients to make trustworthy and clinically justified decisions.

Our findings align with previous research, such as Severi et al., who found high levels of adrenal androgens like androstenedione and DHEA Sulfate associated with a reduced risk of aggressive PC (31). Their study of 17,049 men reported that double concentrations of androstenedione were associated with almost half the risk of aggressive PC, and double concentrations of DHEA-Sulfate were associated with a 37% lower risk. Additionally, Travis et al. described an inverse association of the androstenedione concentration with advanced PC (32).

Our finding of higher SHBG levels in ISUP 3-5 PC complements previous results that proposed SHBG as a marker of aggressive PC in early-stage disease. Preoperative serum SHBG levels were an independent predictor of extraprostatic extension in localized PC, suggesting a role of SHBG in PC progression (33). A study by Price et al. demonstrated that germline genetic variations of androgen-related pathway genes are associated with serum androgen and SHBG concentrations and PC risk (34).

Routine blood test results harbor valuable information that may not be immediately apparent to clinicians. Podnar et al. demonstrated the feasibility of brain tumor diagnosis from routine blood tests using a machine learning predictive model with diagnostic accuracy comparable to imaging studies (35). Regular screening with blood tests for disease prevention is already recommended in many countries (36). This study’s results indicate the potential of simple clinical parameters combined with comprehensive laboratory predictors to estimate aggressive PC risk. Highlighting androgens as complementary diagnostic tools at minimal cost and emphasizing an integrated approach to MRI utilization could lead to more efficient screening strategies for aggressive PC.

Several promising additional biomarkers could complement the current diagnostic pathway for PC, including PSA derivatives and genetic markers (37). Combining the prostate health index and the 4K score (38), if available, with widely accessible and cost-effective clinical and lab values may offer a comprehensive and accurate approach for early non-invasive prediction of aggressive PC. Recognizing the low specificity of PSA as a challenge for PC screening programs, Srivastava et al. recently proposed a “more is more” approach in PC screening programs with the addition of biomarkers in a multistep approach (30). This strategy could help to optimize biopsy decisions, reduce unnecessary biopsies, and improve patient outcomes.

Several limitations must be acknowledged. The pilot study’s sample size may limit the generalizability and statistical power of the findings. Validation in independent, larger cohorts is necessary to ensure the classifier’s robustness and generalizability.

The study population was from a single center, lacking diversity in ethnicity and geographic factors, which may affect the applicability of results to all populations. Identified serum markers, DHEA, SHBG, and androstenedione, need further investigation to validate their efficacy and reliability across diverse populations and their potential for prevention, detection and prognosis of early-stage, aggressive PC. Additionally, the study’s nature did not allow determining causal relationships among variables. Longitudinal studies are necessary to establish relationships between hormone levels and PC development or progression. Lastly, the definition of PC that is aggressive and requires active treatment rather than active surveillance is controversial, with varying definitions of aggressive PC. As ISUP 1-2 PC patients can be offered active surveillance due to the indolent nature of the disease, histological assessment was used to stratify patients into moderate (ISUP 1-2) and aggressive (ISUP 3-5) PC (10,21).

## CONCLUSIONS

Incorporating serum markers DHEA-S, androstenedione, and SHBG into a novel risk classifier can improve early detection of aggressive PC. These widely available and cost-effective blood biomarkers could reduce reliance on invasive prostate biopsies and expensive MRI by providing a more targeted approach to non-invasive prediction of aggressive PC following PSA testing. Our pilot study lays the groundwork for larger-scale research to further explore the integration of androgens and SHBG in future risk stratification models for improved clinical decision making.

## ABBREVIATIONS

AUA: American Urological Association
DHEA: Dehydroepiandrosterone
DRE: Digital rectal examination
EAU: European Association of Urology
ISUP: International Society of Urological Pathology
mpMRI: Multiparametric magnetic resonance imaging
PC: Prostate cancer
PSA: Prostate-specific antigen
PSAD: Prostate-specific antigen density
SHBG: Sex Hormone Binding Globulin

## DECLARATIONS

### Ethical approval

The study was approved by the ethical committee of the University of Heidelberg (Approval No. S-130/2021) and was performed in accordance with the Declaration of Helsinki.

### Contributorship

Conceptualization: M.G.; Data curation: A.R., O.L.; Formal analysis: O.L.; Funding acquisition: M.G., M.H.; Investigation: H.S.; M.G., O.L; Methodology: H.S.; M.G., O.L.; Project Administration: M.G.; Resources: M.G., M.H., O.S.; Software: O.L., O.S.; Supervision: M.G.; Validation: A.R., H.S., O.L.; Visualization: O.L.; Writing - original draft: M.G., O.L.; Writing - review & editing: A.R., H.S., M.H., O.S.

All authors critically reviewed, edited, and added to the manuscript as well as approved the final version of the manuscript.

## Acknowledgements

We sincerely thank Daniela Janscho for her invaluable assistance in handling patient samples.

## Data availability statement

The data sets generated during and analyzed during the current study are available from the corresponding author on reasonable request.

## Declaration of generative AI and AI-assisted technologies in the writing process

During the preparation of this work the authors used ChatGPT-4o in order to improve language and readability. After using this tool, the authors reviewed and edited the content as needed and take full responsibility for the content of the publication.

