## Supplemental Table 1 for "Integration of serum androgens and Sex Hormone-Binding Globulin for optimized early detection of aggressive prostate cancer"

**Supplementary table 1 – Parameters, full p-values and patients’ count**

| **Parameter** | **p-value** | **p-value corrected** | **Type** | **Patients tested** |
| --- | --- | --- | --- | --- |
| Glucose | 0.25750 | 0.51499 | Laboratory measurement | 334 |
| Alkaline Phosphatase | 0.47754 | 0.70374 | Laboratory measurement | 318 |
| Triglycerides | 0.12654 | 0.35432 | Laboratory measurement | 317 |
| HDL Cholesterol | 0.28349 | 0.52918 | Laboratory measurement | 318 |
| C-Reactive Protein | 0.16595 | 0.42242 | Laboratory measurement | 318 |
| Leukocytes | 0.25542 | 0.51499 | Laboratory measurement | 334 |
| Hemoglobin | 0.61595 | 0.74985 | Laboratory measurement | 334 |
| Platelets | 0.91006 | 0.94377 | Laboratory measurement | 334 |
| Neutrophils | 0.59209 | 0.74985 | Laboratory measurement | 317 |
| Absolute Neutrophil Count | 0.33632 | 0.58856 | Laboratory measurement | 316 |
| Absolute Lymphocyte Count | 0.64541 | 0.75297 | Laboratory measurement | 317 |
| Absolute Monocyte Count | 0.01735 | 0.08098 | Laboratory measurement | 316 |
| HbA1c | 0.51004 | 0.71405 | Laboratory measurement | 314 |
| Progesterone | 0.03861 | 0.13934 | Laboratory measurement | 316 |
| Estradiol/E2 | 0.85492 | 0.92068 | Laboratory measurement | 316 |
| Testosterone | 0.79011 | 0.88493 | Laboratory measurement | 315 |
| Androstenedione | 0.00047 | 0.01315 | Laboratory measurement | 320 |
| Dihydrotestosterone | 0.42542 | 0.68826 | Laboratory measurement | 320 |
| PSA | 0.00404 | 0.02963 | Laboratory measurement | 315 |
| Free PSA | 0.03981 | 0.13934 | Laboratory measurement | 315 |
| Free PSA % | 0.00176 | 0.02471 | Laboratory measurement | 315 |
| Vitamin A | 0.08492 | 0.26419 | Laboratory measurement | 315 |
| Vitamin E | 0.95065 | 0.95065 | Laboratory measurement | 315 |
| Zinc (Serum) | 0.19364 | 0.45182 | Laboratory measurement | 313 |
| 25-Hydroxy Vitamin D | 0.57205 | 0.74985 | Laboratory measurement | 315 |
| Cortisol | 0.44245 | 0.68826 | Laboratory measurement | 314 |
| DHEA-Sulfate | 0.00423 | 0.02963 | Laboratory measurement | 315 |
| Sex Hormone Binding Globulin | 0.00652 | 0.03651 | Laboratory measurement | 314 |
| S.p. UTI (last 6 months) | 0.65735 | 0.81603 | Clinical parameter | 553 |
| S.p. urinary retention (last 6 months) | 0.67203 | 0.81603 | Clinical parameter | 553 |
| Meat consumption (days/ week) | 0.89596 | 0.89596 | Clinical parameter | 473 |
| Alcohol (days/week) | 0.38140 | 0.55165 | Clinical parameter | 472 |
| Sport (hours/week) | 0.37537 | 0.55165 | Clinical parameter | 463 |
| Smoking (yes/no) | 0.07088 | 0.20081 | Clinical parameter | 479 |
| Cig. per day | 0.38940 | 0.55165 | Clinical parameter | 501 |
| Smoking (years) | 0.80708 | 0.85752 | Clinical parameter | 478 |
| Age | 0.00000 | 0.00000 | Clinical parameter | 552 |
| BMI | 0.78767 | 0.85752 | Clinical parameter | 485 |
| Positive fam. history for PC | 0.31472 | 0.55165 | Clinical parameter | 490 |
| Infertility | 0.15337 | 0.37246 | Clinical parameter | 553 |
| S.p. sterilization | 0.24981 | 0.53086 | Clinical parameter | 553 |
| Positive DRE | 0.00000 | 0.00000 | Clinical parameter | 553 |
| PSAD | 0.00000 | 0.00000 | Clinical parameter | 314 |
| PIRADS | 0.00000 | 0.00000 | Imaging parameter | 543 |
| Prostate volume | 0.00031 | 0.00105 | Clinical parameter | 552 |
