## Supplemental Table 2 for "Integration of serum androgens and Sex Hormone-Binding Globulin for optimized early detection of aggressive prostate cancer"

**Supplementary table 2 – Median model performance per feature set**

| **Features** | **Accuracy** | **F1** | **Precision** | **Recall** |
| --- | --- | --- | --- | --- |
| **PSA** | 0.44 | 0.39 | 0.35 | 0.448 |
| **+ additional lab. panel** | 0.48 | 0.45 | 0.43 | 0.48 |
| **+ clinical values** | 0.52 | 0.5 | 0.49 | 0.53 |
| **+ MRI results** | 0.56 | 0.54 | 0.56 | 0.56 |
| **MRI + PSA** | 0.53 | 0.51 | 0.55 | 0.53 |
